# Multivariate spatio-temporal analysis of the global COVID-19 pandemic

**DOI:** 10.1101/2021.02.08.21251339

**Authors:** Wen Xiang, Ben Swallow

## Abstract

The COVID-19 pandemic has caused significant mortality and disruption on a global scale not seen in living memory. Understanding the spatial and temporal vectors of transmission as well as similarities in the trajectories of recorded cases and deaths across countries can aid in understanding the benefit or otherwise of varying interventions and control strategies on virus transmission. It can also highlight emerging globa trends as they occur. Data on number of cases and deaths across the globe have been made available through a variety of databases and provide a wide range of opportunities for the application of multivariate statistical methods to extract information on similarity or difference from them. Here we conduct spatial and temporal multivariate statistical analyses of global COVID-19 cases and deaths for the period spanning January to August 2020, using a variety of distance based multivariate methods to cluster countries according to similar temporal trends in cases and deaths resulting from COVID-19. We also use novel air passenger data as a proxy for movement between countries. The air passenger movement can act as an important vector of transmission and thus scaling covariance matrices before conducting dimension reduction techniques can account for known structures in the data and help highlight important residual spatial and/or temporal trends that may then be attributable to the success of interventions or other cultural differences. Global temporal structure is found to be of significantly more importance than local spatial structure in terms of global dynamics. Our results highlight a significant global change in case and mortality daynamics from early-August, consistent in timing with the emergence of new strains with highger levels of transmission. We propose the methodology offers great potential in real-time analysis of complex, noisy spatio-temporal data and the extraction of emerging changes in pandemic dynamics that can support policy and decision makers.

## 1 Introduction

The SARS-COV-2 pandemic is a global crisis not seen on such a scale in peace-time living memory, yet little has so far been considered of the spatial and temporal dynamics of the pandemic on a global scale, despite strong reasons for doing so^1^. Whilst the virus knows nothing of the political boundaries of geographical regions, the interventions taken to hinder the virus’ progression have varied significantly across countries and adherence to these interventions can vary drastically depending on.^2^ Understanding the important similarities in the dynamics of the pandemic across both space and time is therefore of huge importance in aiding successful policy interventions.

Multivariate methods are a collection of statistical methods and computational algorithms for analysing data where multiple measurements are taken on each sampled unit. Among these, clustering methods aim to join together units that are similar to each other in the same group, with units that are inherently different placed in different groups.^3^ Several approaches have extended standard clustering methods to account for the inherent correlation present in time-series of observations^4^, and therefore cluster complete temporal trajectories rather than single observations. The ability to cluster time series data offers the potential to study how trajectories of observed cases and/or deaths attributed to COVID-19 vary across countries and continents, ideally accounting for the fact that emergence time and duration of regional epidemics will vary, and hence time series could be of equal or unequal length.

In addition to this, multivariate projection and decomposition methods, such as Principal Components Analysis (PCA),^5^ allow extraction of important structures inherent in multivariate data through an eigen-decomposition of the correlation or covariance matrix. However, in general these unsepervised methods find lower-dimensional subspaces in the data without explicitly using known structures and are based on an unaided orthogonal, variance maximisation approach. Extensions to PCA relax this assumption of unknown structure and allow users to account for existing spatial and/or temporal structures inherent in the data through the use of spatial and/or temporal weighting matrices.^7^ Spatial and temporal weight matrices have been previously used to account for important flow vectors in geostatistics, specifically in river flow networks, and incorporated into existing dimension reduction methodologies. Accounting for existent spatial and temporal structures in the data allow the extraction of important residual joint structures that can be more readily interpreted than if these known structures are not accounted for. Using data on air passenger flow between airports in impacted countries, we treat the flow of people between countries in a similar manner to hydrological flow, and then ascribe additional remaining temporal and spatial structure to the efficacy of the variety of interventions that have been implemented on a global scale.

In addition to the potential transmission risk between passengers onboard planes,^8^ travel of infected patients between countries is seen as an important vector of transmission before large-scale interventions were introduced.^10^ Air travel is also a likely proxy of more general traffic between nations. The possible impact of those interventions, as well as testing for similarities in space and time between countries with different political, social and economic standing, is therefore of great interest.

Using ECDC reported case and death data between 2020-01-01 and 2020-08-22, normalised by population size in each country^11^, we use a variety of algorithmic clustering and multivariate dimension reduction methods to cluster countries according to similar spatial and/or temporal trends in cases and deaths attributed to COVID-19 or help detect important change points in global pandemic dynamics.

## 2 Results

### 2.1 Clustering based on ACF

We initially study the temporal dynamics of COVID-19 reported case and death numbers using algorithmic fuzzy clustering to join together similar time series across countries. Persistent autocorrelation at increasing lags up to 20 weeks highlights countries where cases remain consistently high or low for long periods of time. Conversely, sharply decreasing autocorrelation would imply that observed numbers are relatively disctinct over all but the shortest periods. The 210 countries were allocated into one of six clusters with similar temporal dynamics in case numbers (Figure 1 and Table 1) and deaths (Figure 2 and Table 2). Clustering was conducted using dynamic time warping (henceforth DTW) on the autocorrelation lag series of increasing lengths and on the underlying per capita time series.

**Table 1:** Cluster members for cases.

| Cluster_1 | Cluster_2 | Cluster_3 | Cluster_4 | Cluster_5 | Cluster_6 |
| --- | --- | --- | --- | --- | --- |
| Andorra | Afghanistan | Argentina | Anguilla | Barbados | Austria |
| Angola | Albania | Australia | Antigua and Barbuda | Burkina Faso | Bahamas |
| Aruba | Algeria | Belgium | Benin | Cases on an international conveyance Japan | Cape Verde |
| Belize | Armenia | Bosnia and Herzegovina | Bermuda | Cayman Islands | Central African Republic |
| Brunei Darussalam | Azerbaijan | Bulgaria | Bhutan | Congo | Cote d'Ivoire |
| Chad | Bahrain | Costa Rica | Bonaire, Saint Eustatius and Saba | Djibouti | Croatia |
| Chile | Bangladesh | Czechia | Botswana | Ecuador | Cuba |
| China | Belarus | Denmark | British Virgin Islands | Faroe Islands | Cyprus |
| Gambia | Bolivia | Eswatini | Burundi | Gabon | Democratic Republic of the Congo |
| Georgia | Brazil | Finland | Cambodia | Gibraltar | Estonia |
| Ghana | Canada | Germany | Cameroon | Guam | Ethiopia |
| Guernsey | Colombia | Iceland | Comoros | Guinea | France |
| Guyana | Dominican Republic | Ireland | Curaçao | Guinea Bissau | Greece |
| Isle of Man | Egypt | Japan | Dominica | Jamaica | Guatemala |
| Kosovo | El Salvador | Madagascar | Equatorial Guinea | Jersey | Haiti |
| Latvia | Honduras | Malaysia | Eritrea | Jordan | Hungary |
| Liberia | India | North Macedonia | Falkland Islands (Malvinas) | Kazakhstan | Lebanon |
| Libya | Indonesia | Norway | Fiji | Kyrgyzstan | Lithuania |
| Malta | Iran | Peru | French Polynesia | Laos | Luxembourg |
| Mauritius | Iraq | Philippines | Greenland | Lesotho | Malawi |
| Monaco | Israel | Senegal | Grenada | Liechtenstein | Maldives |
| Montenegro | Italy | Serbia | Holy See | Mali | Moldova |
| Niger | Kenya | Singapore | Mongolia | Mauritania | Morocco |
| Portugal | Kuwait | Suriname | Montserrat | New Caledonia | Mozambique |
| Republic of Korea | Mexico | Switzerland | Myanmar | Papua New Guinea | Namibia |
| Rwanda | Netherlands | Turkey | Nicaragua | Sri Lanka | Nepal |
| San Marino | Nigeria | - | Northern Mariana Islands | Tajikistan | New Zealand |
| Sierra Leone | Oman | - | Saint Kitts and Nevis | Togo | Paraguay |
| Sint Maarten | Pakistan | - | Saint Lucia | Tunisia | Poland |
| Slovakia | Palestine | - | Saint Vincent and the Grenadines | Uganda | Puerto Rico |
| Somalia | Panama | - | Sao Tome and Principe | Uruguay | Slovenia |
| Sudan | Qatar | - | Seychelles | Yemen | Spain |
| Syria | Romania | - | South Sudan | - | Sweden |
| Taiwan | Russia | - | Timor Leste | - | Thailand |
| Trinidad and Tobago | Saudi Arabia | - | United Republic of Tanzania | - | Ukraine |
| Turks and Caicos islands | South Africa | - | Western Sahara | - | Venezuela |
| United States Virgin Islands | United Arab Emirates | - | - | - | Zambia |
| Vietnam | United Kingdom | - | - | - | Zimbabwe |
| - | United States | - | - | - | - |
| - | Uzbekistan | - | - | - | - |

**Table 2:** Cluster members for deaths using autocorrelation lag.

| Cluster_1 | Cluster_2 | Cluster_3 | Cluster_4 | Cluster_5 | Cluster_6 |
| --- | --- | --- | --- | --- | --- |
| Andorra | Algeria | Bahamas | Afghanistan | Armenia | Anguilla |
| Angola | Antigua and Barbuda | Barbados | Albania | Azerbaijan | Bhutan |
| Bosnia and Herzegovina | Aruba | Benin | Argentina | Bangladesh | Bonaire, Saint Eustatius and Saba |
| Bulgaria | Belize | Burkina Faso | Australia | Belarus | Cambodia |
| Croatia | Bermuda | Cape Verde | Austria | Belgium | Dominica |
| Cuba | Botswana | Central African Republic | Bahrain | Bolivia | Eritrea |
| El Salvador | British Virgin Islands | Chad | China | Brazil | Falkland Islands (Malvinas) |
| Estonia | Brunei Darussalam | Chile | Czechia | Canada | Faroe Islands |
| Eswatini | Burundi | Cote d'Ivoire | Dominican Republic | Colombia | French Polynesia |
| Finland | Cameroon | Democratic Republic of the Congo | Ethiopia | Costa Rica | Gibraltar |
| Greece | Cases on an international conveyance Japan | Djibouti | France | Denmark | Greenland |
| Honduras | Cayman Islands | Ecuador | Guatemala | Egypt | Grenada |
| Ireland | Comoros | Gambia | Hungary | Germany | Holy See |
| Israel | Congo | Ghana | Madagascar | India | Laos |
| Japan | Curacao | Guernsey | Malaysia | Indonesia | Mongolia |
| Kenya | Cyprus | Haiti | Nigeria | Iran | New Caledonia |
| Lebanon | Equatorial Guinea | Iceland | North Macedonia | Iraq | Saint Kitts and Nevis |
| Libya | Fiji | Isle of Man | Norway | Italy | Saint Lucia |
| Luxembourg | Gabon | Jersey | Portugal | Kuwait | Saint Vincent and the Grenadines |
| Malawi | Georgia | Kosovo | Qatar | Mexico | Seychelles |
| Moldova | Guam | Lesotho | Republic of Korea | Netherlands | Timor Leste |
| Montenegro | Guinea | Lithuania | Serbia | Pakistan | - |
| Morocco | Guinea Bissau | Mali | Spain | Panama | - |
| Namibia | Guyana | Mauritania | Sweden | Romania | - |
| Nepal | Jamaica | New Zealand | Switzerland | Russia | - |
| Oman | Jordan | Niger | United Arab Emirates | Saudi Arabia | - |
| Palestine | Kazakhstan | Philippines | Venezuela | South Africa | - |
| Paraguay | Kyrgyzstan | San Marino | - | Turkey | - |
| Poland | Latvia | Sierra Leone | - | United Kingdom | - |
| Puerto Rico | Liberia | Singapore | - | United States | - |
| Senegal | Liechtenstein | Slovakia | - | Uzbekistan | - |
| Slovenia | Maldives | Somalia | - | - | - |
| Sudan | Malta | Suriname | - | - | - |
| Thailand | Mauritius | Syria | - | - | - |
| Ukraine | Monaco | Tajikistan | - | - | - |
| Zimbabwe | Montserrat | Tunisia | - | - | - |
| - | Mozambique | Uganda | - | - | - |
| - | Myanmar | Vietnam | - | - | - |
| - | Nicaragua | Yemen | - | - | - |
| - | Northern Mariana Islands | Zambia | - | - | - |
| - | Papua New Guinea | - | - | - | - |
| - | Peru | - | - | - | - |
| - | Rwanda | - | - | - | - |
| - | Sao Tome and Principe | - | - | - | - |
| - | Sint Maarten | - | - | - | - |
| - | South Sudan | - | - | - | - |
| - | Sri Lanka | - | - | - | - |
| - | Taiwan | - | - | - | - |
| - | Togo | - | - | - | - |
| - | Trinidad and Tobago | - | - | - | - |
| - | Turks and Caicos Islands | - | - | - | - |
| - | United Republic of Tanzania | - | - | - | - |
| - | United States Virgin Islands | - | - | - | - |
| - | Uruguay | - | - | - | - |
| - | Western Sahara | - | - | - | - |

**Figure 1:**
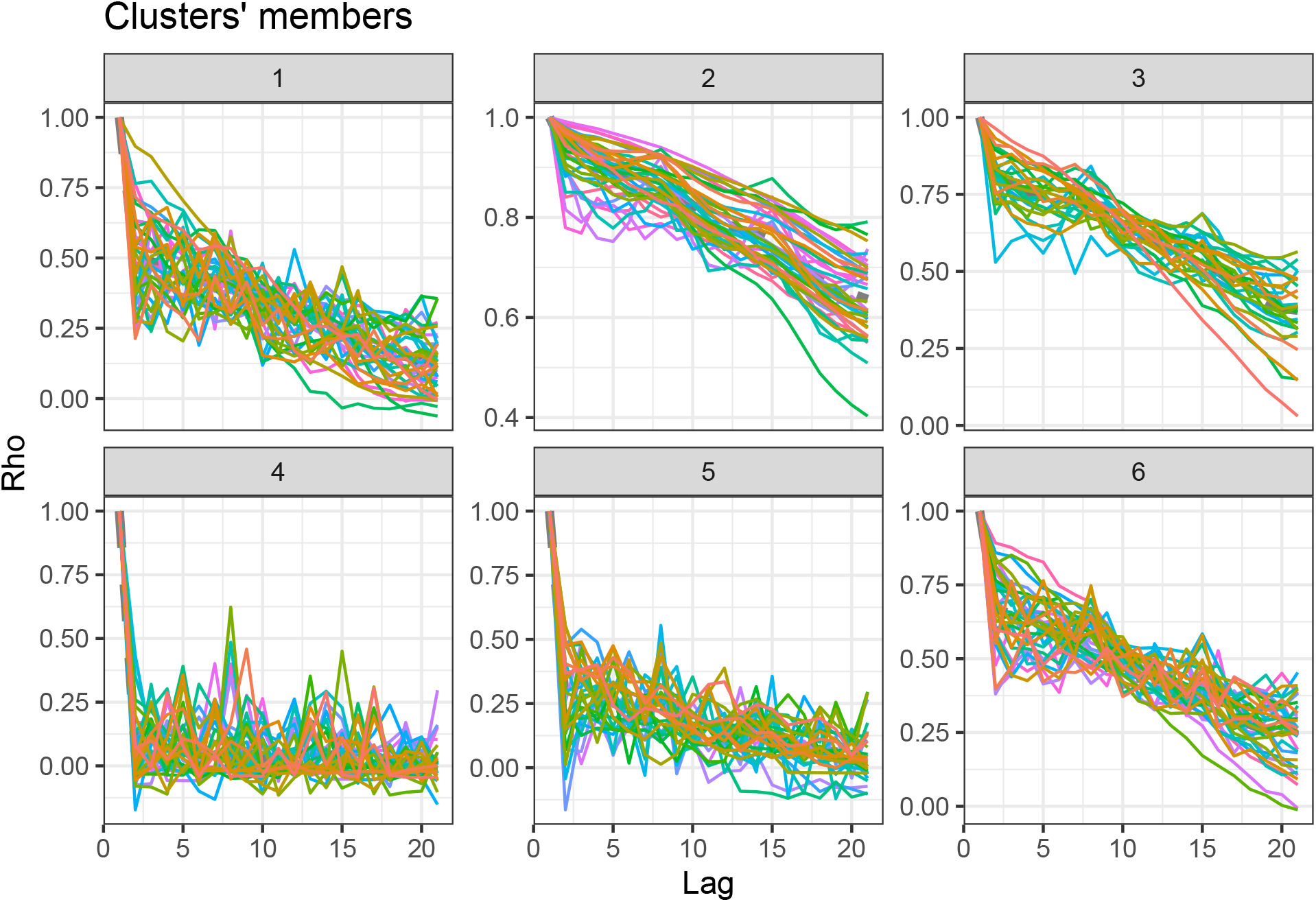
Temporal lag (x-axis) plotted against correlation coefficient for the *k* = 6 clusters.

**Figure 2:**
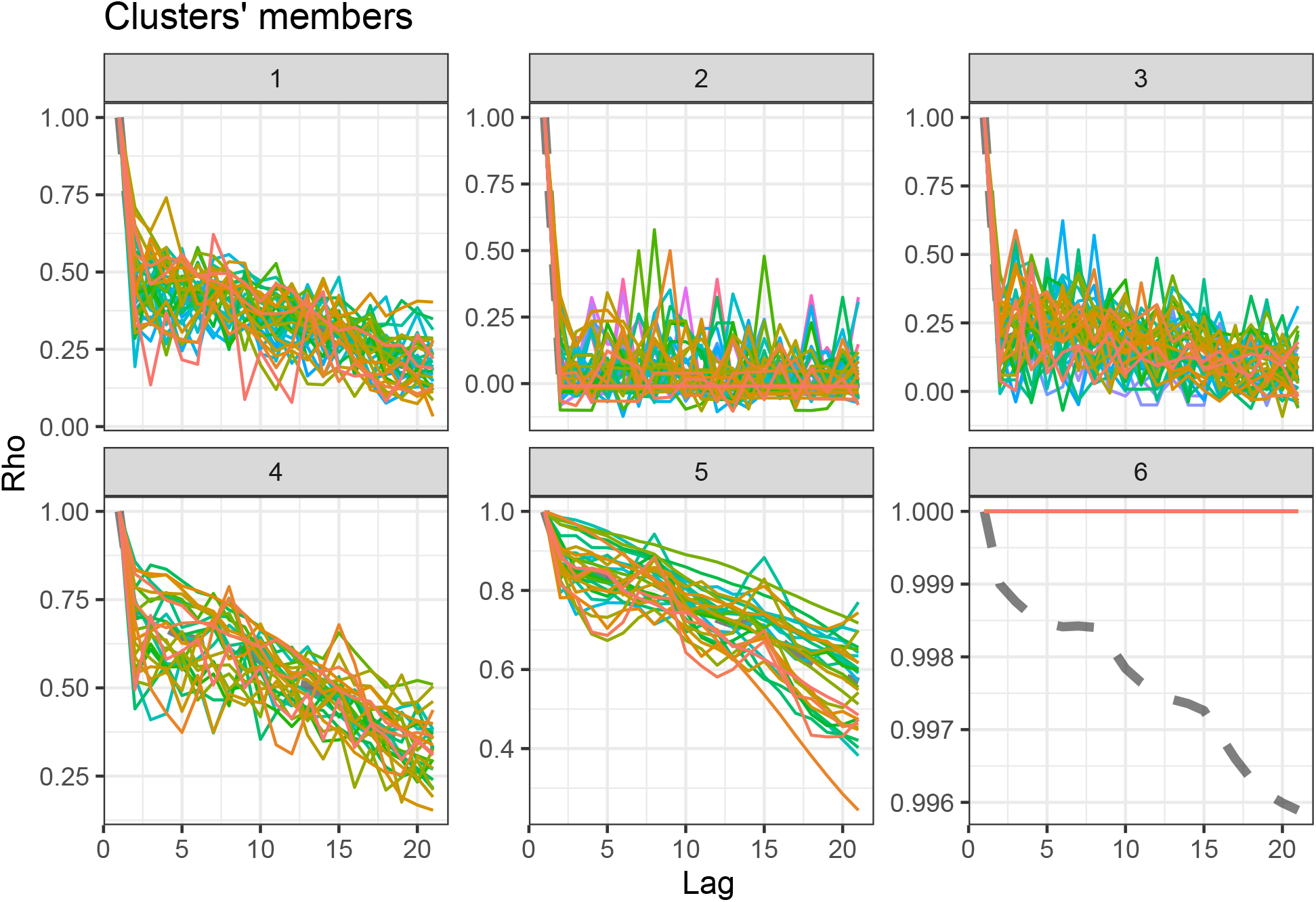
Temporal lag (x-axis) plotted against correlation coefficient for number of recorded deaths.

#### 2.1.1 Cases

Multiple different structures across clusterings can be observed. Specifically, clusters 2 and 3 correspond to countries with very shallow decreases in autocorrelation. This is particularly strong in cluster 2, where autocorrelation is still greater than 0.6 at lag 20 for almost all of the cluster members. Cluster 4 corresponds to countries with very minimal autocorrelation even at the lowest lags. This cluster contains countries with little consistency in observed cases or large degrees of fluctuation over time. Clusters 1 and 5 on the other hand, show moderate correlation at low lags, with a relatively steep reduction towards lag 20. These countries are showing short-term temporal dependence but again cases drop off swiftly. Finally cluster 6 appears to show countries for which, whilst there is significant dependence on a short-term basis, this very swiftly drops down towards zero by 20 days later.

#### 2.1.2 Deaths

Similar types of structures could be observed in the mortality data, although the countries exhibiting these trends varied. The main difference was the countries in cluster 6, which showed almost consistent autocorrelation of one up to and including the highest lag. These countries show little variation in numbers of deaths, mostly due to very low figures.

Adjusted Rand index between the two clusters was 0.23, suggesting a relatively low level of similarity between the two sets of clusters and therefore differing factors underlying the temporal trajectories.

### 2.2 Clustering based on distance measures between timeseries

We can also use distance measures on the original time-series (again DTW) but this only works for the time periods where there is concurrent complete data across all countries, as the series must be of the same length. Number of cases/deaths are plotted against day index, with dates ranging from 2020-05-17 to 2020-08-23.

#### 2.2.1 Cases

Figure 3 shows the time series of cases per capita in each of the 12 clusters, with the corresponding cluster members in Table 3. The method is capable of clustering similar trajectories and magnitudes, even with shifts in time between the dynamics.

**Table 3:**
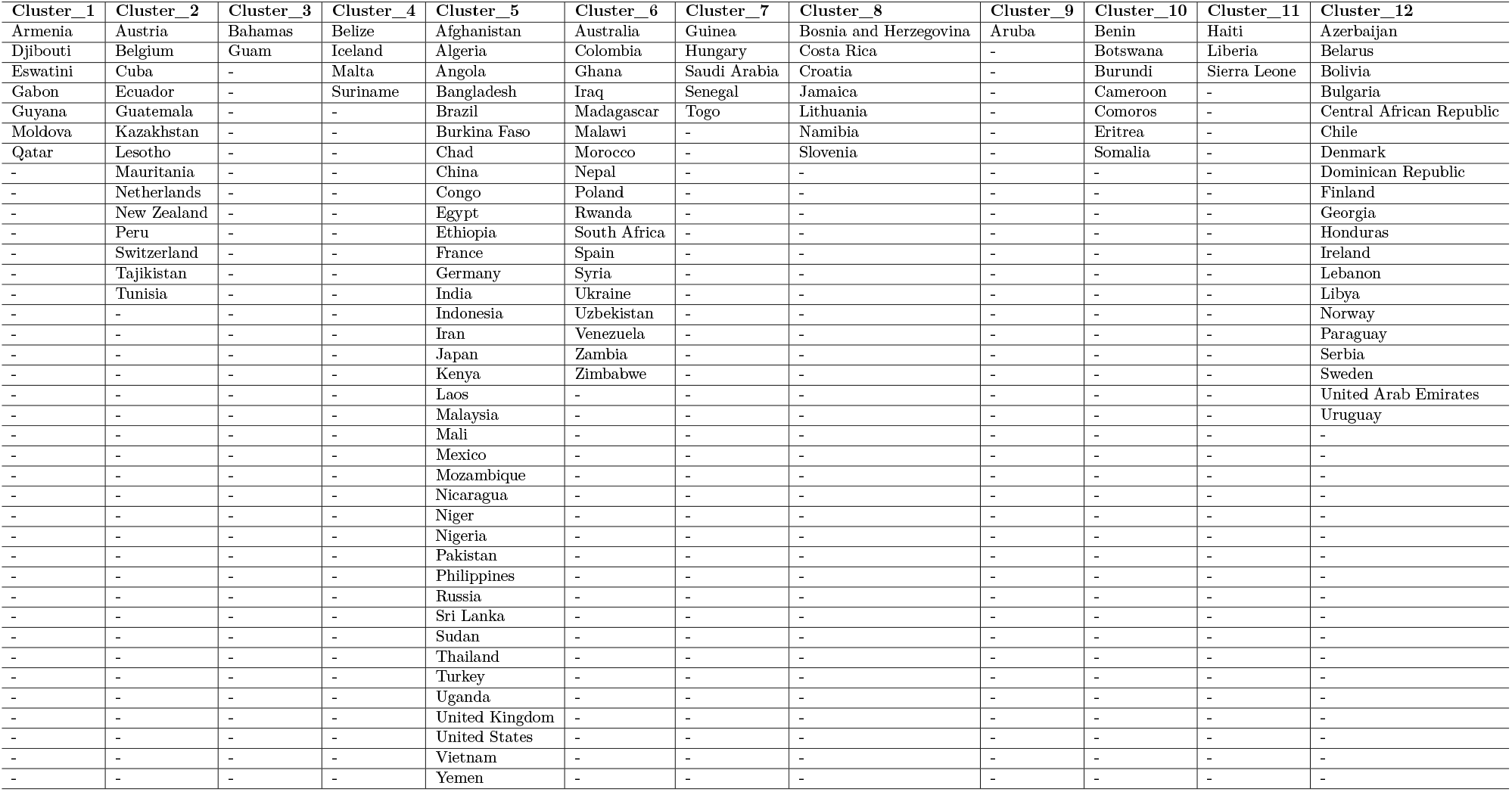
Cluster members for cases using distance between timeseries.

**Figure 3:**
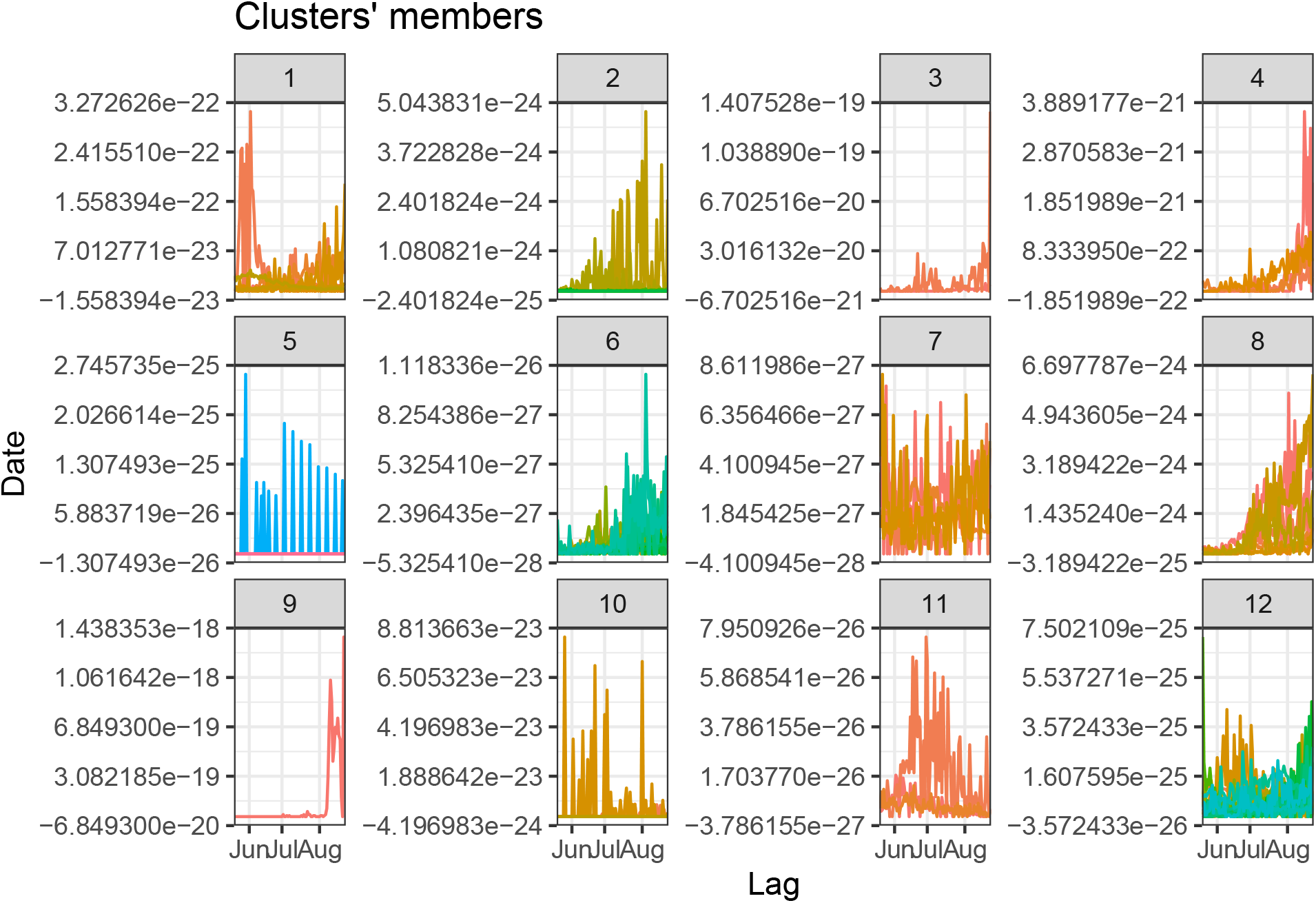
Date in 2020 (x-axis) plotted against cases per capita.

#### 2.2.2 Deaths

Figure 4 shows the time series of deaths per capita in each of the 12 clusters, with the corresponding cluster members in Table 4. Once again the clusters contain time series of similar dynamics and magnitude.

**Table 4:** Cluster members for deaths using distance between timeseries.

| Cluster_1 | Cluster_2 | Cluster_3 | Cluster_4 | Cluster_5 | Cluster_6 | Cluster_7 | Cluster_8 | Cluster_9 | Cluster_10 | Cluster_11 | Cluster_12 |
| --- | --- | --- | --- | --- | --- | --- | --- | --- | --- | --- | --- |
| Bahamas | Central African Republic | Armenia | Azerbaijan | Aruba | Bosnia and Herzegovina | Algeria | Austria | Bolivia | Brazil | Afghanistan | Australia |
| Bangladesh | Croatia | Djibouti | Belarus | - | Lithuania | Iran | Belgium | Bulgaria | Egypt | Angola | Colombia |
| Belize | Denmark | Eswatini | Chile | - | Mauritania | Mexico | Ecuador | Costa Rica | France | Benin | Haiti |
| Botswana | Finland | Gabon | Dominican Republic | - | Namibia | Morocco | Hungary | Honduras | Germany | Burkina Faso | Iraq |
| Burundi | Ireland | Guyana | Guatemala | - | Qatar | Nepal | Netherlands | Libya | Kenya | Cameroon | Kazakhstan |
| China | Liberia | Lesotho | - | - | - | Poland | Peru | Paraguay | Russia | Chad | Lebanon |
| Congo | Norway | Moldova | - | - | - | Ukraine | Sierra Leone | Serbia | Spain | Comoros | Malawi |
| Eritrea | Sweden | Slovenia | - | - | - | Uzbekistan | United Arab Emirates | - | Turkey | Cuba | Saudi Arabia |
| Ethiopia | Uruguay | Suriname | - | - | - | Venezuela | - | - | - | Ghana | Senegal |
| Georgia | - | - | - | - | - | Yemen | - | - | - | Guinea | South Africa |
| Guam | - | - | - | - | - | - | - | - | - | Madagascar | Zambia |
| Iceland | - | - | - | - | - | - | - | - | - | Mali | Zimbabwe |
| India | - | - | - | - | - | - | - | - | - | Niger | - |
| Indonesia | - | - | - | - | - | - | - | - | - | Rwanda | - |
| Jamaica | - | - | - | - | - | - | - | - | - | Somalia | - |
| Japan | - | - | - | - | - | - | - | - | - | Sudan | - |
| Laos | - | - | - | - | - | - | - | - | - | Switzerland | - |
| Malaysia | - | - | - | - | - | - | - | - | - | Syria | - |
| Malta | - | - | - | - | - | - | - | - | - | Togo | - |
| Mozambique | - | - | - | - | - | - | - | - | - | Tunisia | - |
| New Zealand | - | - | - | - | - | - | - | - | - | United Kingdom | - |
| Nicaragua | - | - | - | - | - | - | - | - | - | - | - |
| Nigeria | - | - | - | - | - | - | - | - | - | - | - |
| Pakistan | - | - | - | - | - | - | - | - | - | - | - |
| Philippines | - | - | - | - | - | - | - | - | - | - | - |
| Sri Lanka | - | - | - | - | - | - | - | - | - | - | - |
| Tajikistan | - | - | - | - | - | - | - | - | - | - | - |
| Thailand | - | - | - | - | - | - | - | - | - | - | - |
| Uganda | - | - | - | - | - | - | - | - | - | - | - |
| United States | - | - | - | - | - | - | - | - | - | - | - |
| Vietnam | - | - | - | - | - | - | - | - | - | - | - |

**Figure 4:**
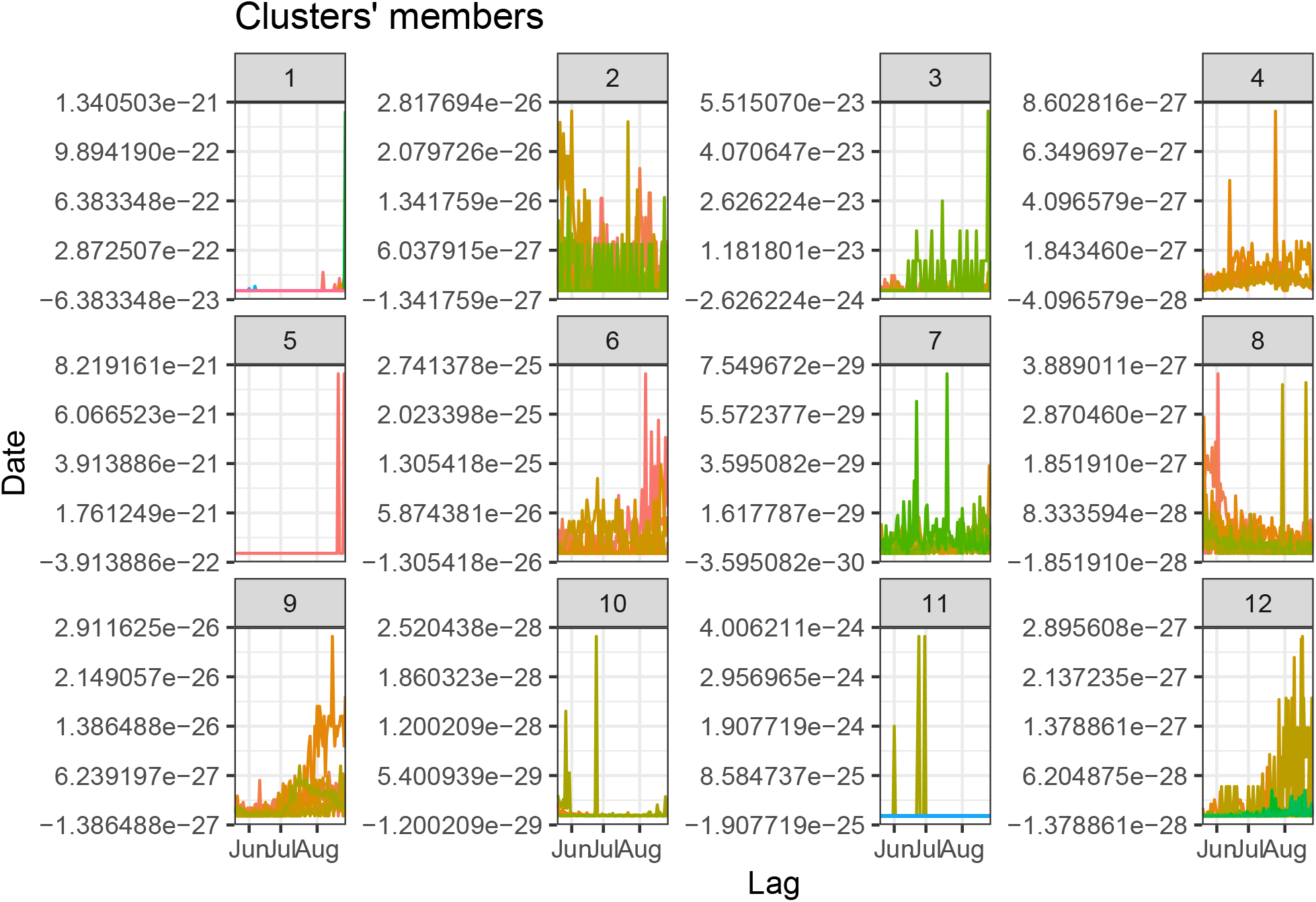
Date in 2020 (x-axis) plotted against deaths per capita.

Adjusted Rand index between the two clusters was 0.22 again suggesting a relatively low level of similarity between the two sets of clusters.

### 2.3 Flow directed PCA

Our second approach to determining dynamics is to conduct variants of PCA to produce low-dimensional representations of the data matrix.

All analyses were dominated by the first principal component, which explained approximately 99% of variation in all settings. Also note that the dimension reduction is identifiable up to a sign change, so some of the scores plots show similar dynamics but inverted because of this non-identifiability.

#### 2.3.1 S-Mode - cases

The first three principal scores from the S-Mode analyses are presented in Figure 5 with corresponding proportions of variance explained in Table 5. The analysis of case data shows a relatively consistent dominant trend across all three weighting procedures, with the first component accounting for around 99% of variability. This suggests very little spatial variability in temporal dynamics.^7^ Relatively little consistency across countries is shown between mid-May and, with a steep global spike in cases observed from mid-August onwards. This suggests that after normalising by population size, there was no strong temporal trend in global case numbers until mid-August, when there was a sudden sharp increase in the total global population of infected individuals. The second principal score had strongest contribution to overall variance explanation in the spatio-temporal analysis. This less-important trend shows an additional minimal peak in the second half of June, and a further large spike again from the second week of August onwards. The second spike is particularly pronounced after removing country-specific temporal variation.

**Table 5:** Percentage of case per capita variance explained by S-mode PC1, PC2 and PC3 for the three different weight matrics.

| Unweighted | Spatial | Spatiotemporal |
| --- | --- | --- |
| 99.7 | 99.7 | 99.27 |
| 0.3 | 0.3 | 0.73 |
| 0.0 | 0.0 | 0.00 |

**Figure 5:**
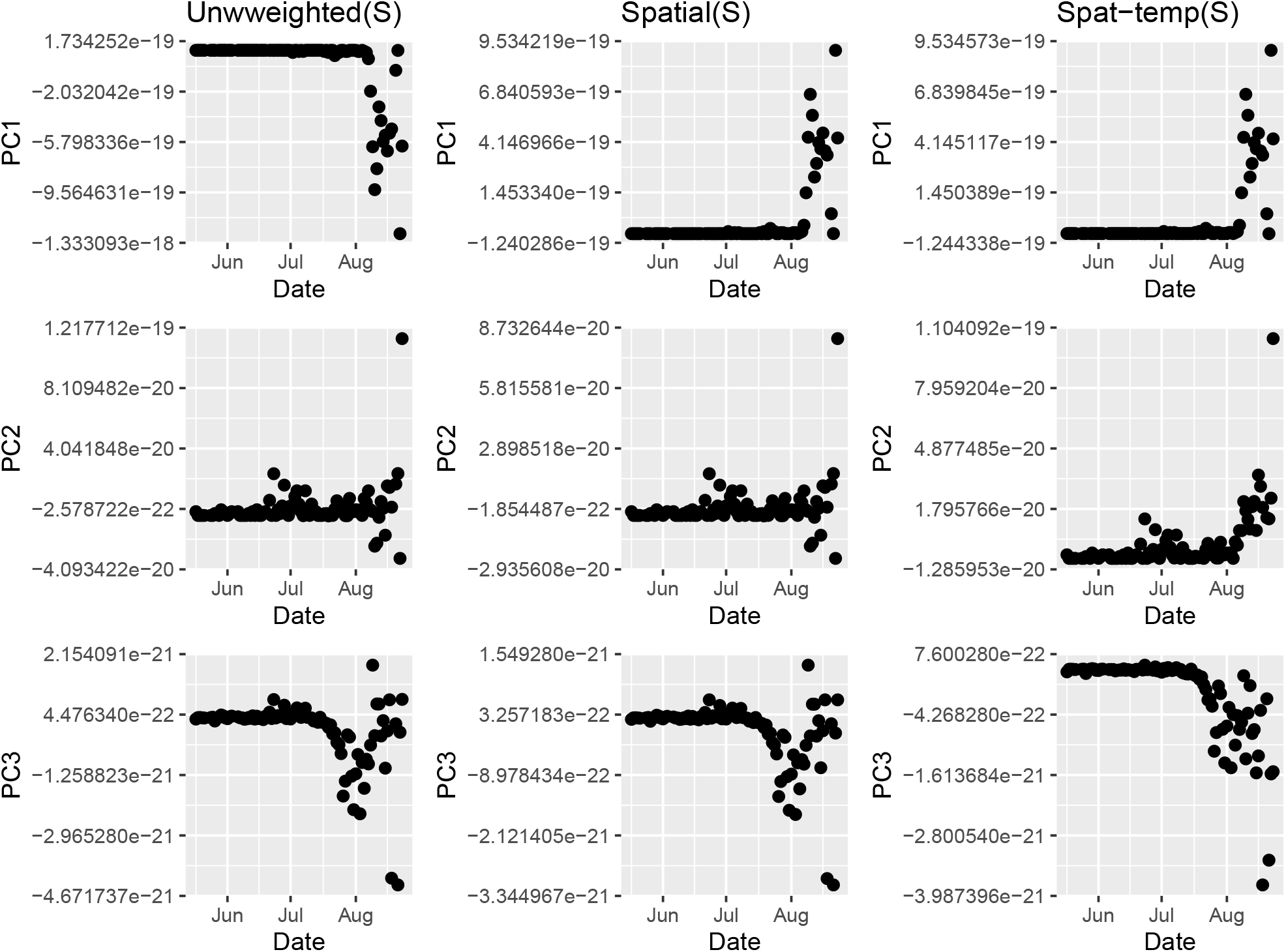
First three weighted S-Mode PCA scores on number of cases with different weight matricies applied.

#### 2.3.2 S-Mode - deaths

The results from the S-Mode analysis are presented in Figure 6 with corresponding proportions of variance explained in Table 6. The analysis of deaths proved problematic, due to obvious discrepancies in the data. There were significantly large values of deaths in several of the later weeks that were not specific to a single country and these dominated the first two principal components, as can be seen in Figure. Given the fact that these involved multiple countries simultaneously, it is hard to justify removing them completely. Looking at the third component, however, there appears to be some additional noticeable change in deaths approximately a week after the emergence of the spike observed in the analysis of cases. This will likely represent more relevant variation in mortality data similar to that detected in the cases data. Given the discrepancy in time between the two spikes, it suggests that observed increases in deaths occurred a week later. This seems reasonable given that mean time from illness onset to death has been estimated as between 15.1 and 29.5,^12^ but this would need to include time for formal testing and reporting of cases (estimated at 7.1 days).^12^

**Table 6:** Percentage of death per capita variance explained by S-mode PC1, PC2 and PC3 for the three different weight matrics.

| Unweighted | Spatial | Spatiotemporal |
| --- | --- | --- |
| 99.37 | 99.37 | 98.99 |
| 0.60 | 0.61 | 0.97 |
| 0.02 | 0.02 | 0.03 |

**Figure 6:**
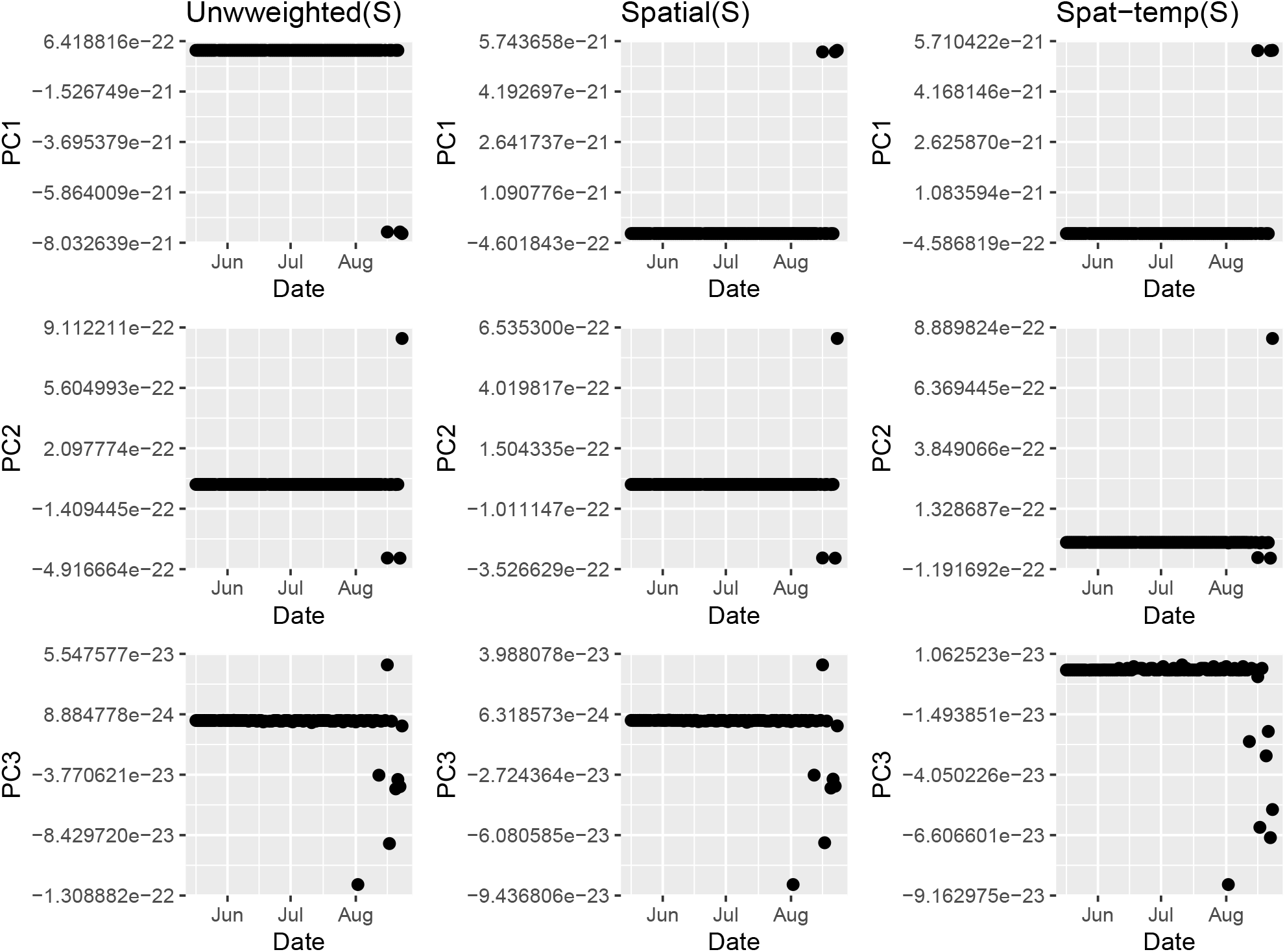
First three weighted S-Mode PCA scores on number of deaths with different weight matricies applied.

#### 2.3.3 T-Mode - cases

The results from the T-Mode analysis of cases are presented in Figure 7 with corresponding proportions of variance explained in Table 7. The results are a stark contrast to the S-Mode analyses is showing very little spatial structure. The only nation to be highlighted in the first PC is Aruba, which is shown to have a significantly different case-per-capita temporal series than other countries.

**Table 7:** Percentage of case per capita variance explained by T-mode PC1, PC2 and PC3 for the three different weight matrics.

| Unweighted | Spatial | Spatiotemporal |
| --- | --- | --- |
| 99.72 | 99.72 | 99.26 |
| 0.28 | 0.28 | 0.74 |
| 0.00 | 0.00 | 0.00 |

**Figure 7:**
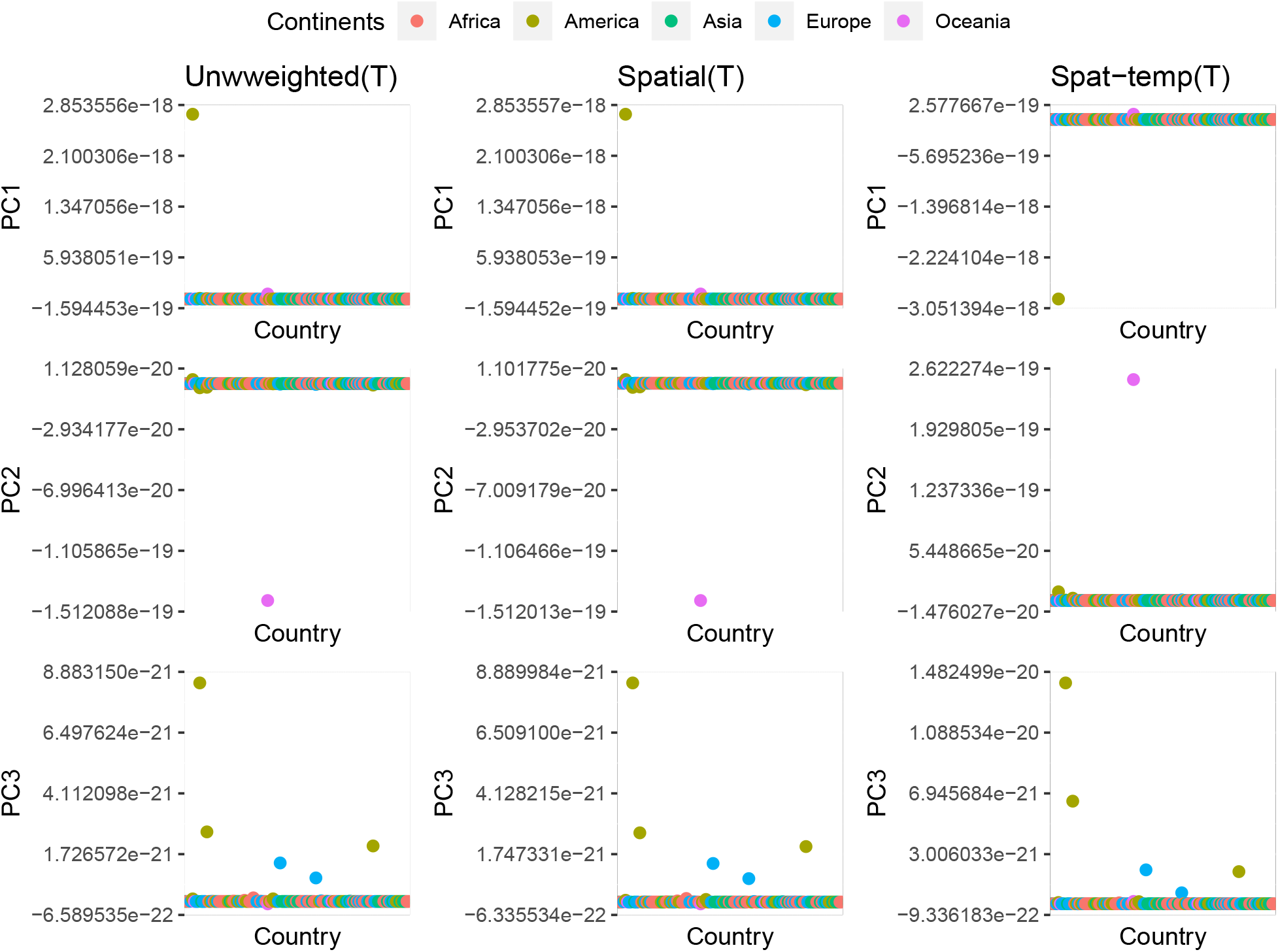
Spatio-temporally weighted T-Mode PCA scores on number of cases (plotted alphabetically).

#### 2.3.4 T-Mode - deaths

The results from the T-Mode analysis of deaths are presented in Figures 8 with corresponding proportions of variance explained in Table 8. Once again, very little consistent spatial structure was observed in the data, with Aruba being the single country highlighted by the analysis as having a particularly unique deaths per capita trend.

**Table 8:** Percentage of death per capita variance explained by T-mode PC1, PC2 and PC3 for the three different weight matrics.

| Unweighted | Spatial | Spatiotemporal |
| --- | --- | --- |
| 99.39 | 99.38 | 98.99 |
| 0.59 | 0.59 | 0.97 |
| 0.02 | 0.02 | 0.03 |

**Figure 8:**
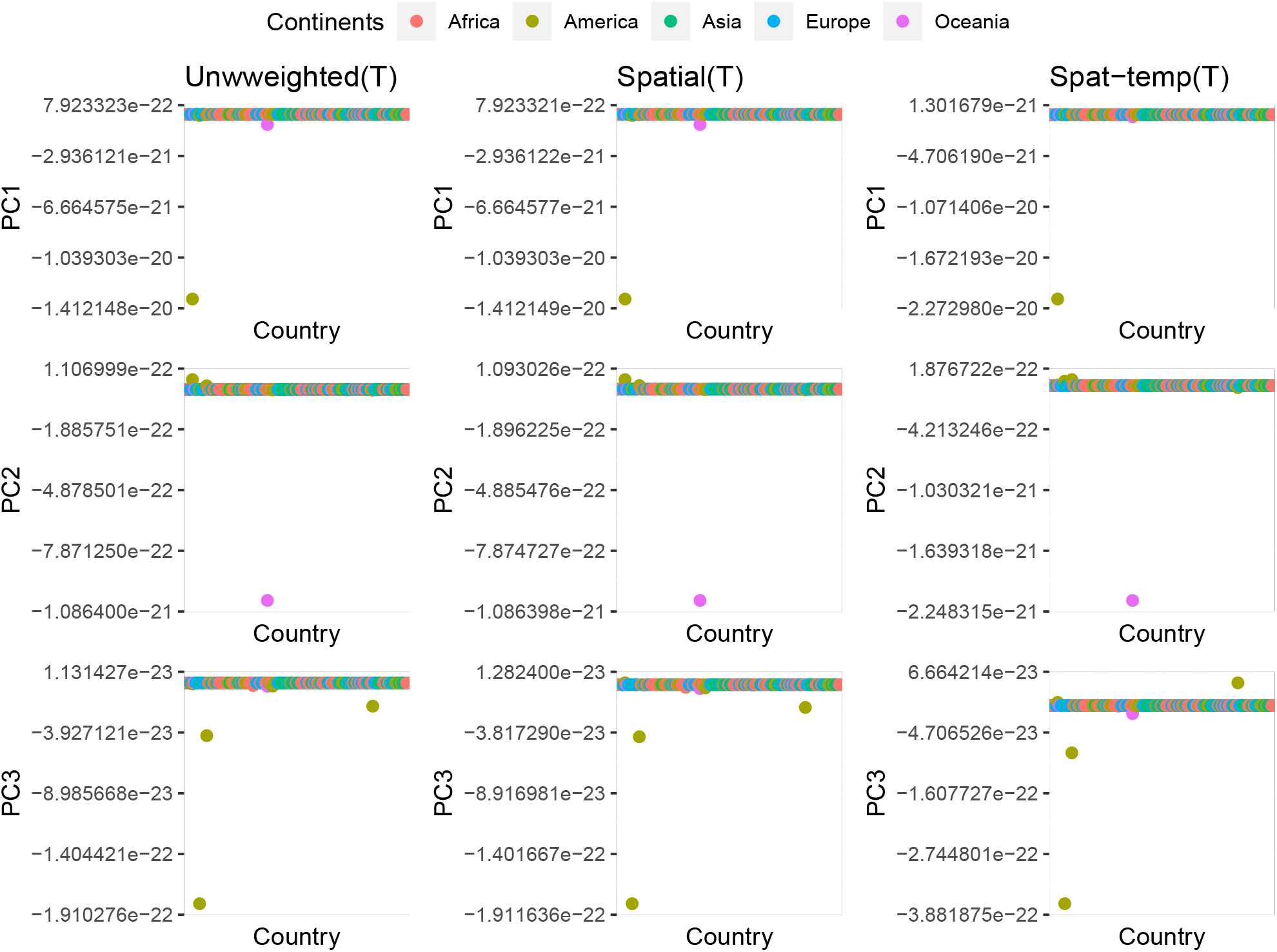
Spatio-temporally weighted T-Mode PCA scores on number of deaths (plotted alphabetically).

## 3 Methods

We employ two broad approaches to detecting dynamics across space and time using several publicly available datasets, initially using algorithmic temporal clustering methods to cluster countries according to different measures of temporal (dis)similarity. Our second approach is to study spatial and temporal structure through weighted PCA methodology. All statistical analyses were conducted in R version 4.0.2.

### 3.1 Data

Daily case and incidence data were extracted from.^13^ Dates were converted to ‘days since 2020-01-01’ for ease of modelling. The number of cases for Iran were not added on 2020-04-04 and approximately double were added on 2020-04-05, so these were shared equally across the two dates. An unrealistically high value of deaths were also observed for China on 2020-04-17 which corresponded to historical update of records that had been previously attributed to other diagnoses,^14^ and hence this datum was removed.

Global population data dating from 2019 was extracted from,^11^ with countries matched to those in the incidence data. Data for Eritrea was only available up until 2011, so this figure was used instead for consistency with the other country methodologies. Case and deaths for each country were divided by these population figures to give incidence per capita values.

### 3.2 Clustering on temporal lags

We initially cluster countries based on the autocorrelation profile^15^, namely calculating autocorrelation up to lag 20 for each country. DTW is a method for calculating the optimum match between two series, allowing for the fact that different countries may have similar trajectories but displaced in time depending on the onset of emergence. With this approach, we can use the complete data trajectories by clustering based on the increasing lags over the course of the pandemic, even when the time series differ in length. Clusters are calculated based on a fuzzy clustering approach^16^ with L2-norm distance between autocorrelation functions, allowing calculation of probabilities associated with belonging to each cluster. Countries are assigned the cluster with highest probability.

Temporal clustering was carried out using various functions in the dtwclust package in R^15^. Dynamic Time Warping^17^ is an algorithm that calculates distance between two temporal series that may be of different lengths, aiming to calculate an optimal match between the indices of two given sequences.

### 3.3 Clustering on incidence per capita

We also cluster in a more traditional way according to either the number of cases or deaths per capita. In this case, all time series must be of the same length so only the 126 countries with complete data during a core central period are included. Again using dynamic time warping as the distance metric, we employ a k-medoids partitioning algorithm with 12 clusters to form clusters of similar trajectories in cases and deaths^15^. A larger number of clusters were used to account for greater variability in these raw data.

In both of the above scenarios, we use the Adjusted Rand Index (ARI)^18^ to compare between clusterings of cases (denoted *X*_*i*_) and deaths (denoted *Y*_*j*_) under each of the two approaches. The ARI is defined as

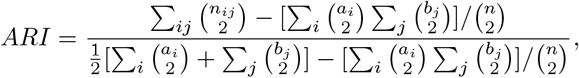

where *n*_*i,j*_ denotes the number of objects in common between cluster *X*_*i*_ and *Y*_*j*_, *a*_*i*_ = Σ_*j*_ *n*_*i,j*_ and *b*_*j*_ = Σ_*i*_ *n*_*i,j*_. Values close to one suggest similar cluster memberships and values close to zero denote very different clusters. Due to the lower number of countries in the latter setting, it is not feasible to compare clusterings between methods.

### 3.4 Flow directed PCA

Next we aim to discover important global spatial and temporal trends in cases and deaths from COVID-19. To extract the important trends, PCA and similar dimension reduction techniques are an obvious choice. PCA conducts an eigendecomposition of the covariance (or correlation) of a data matrix, with eigenvalues ordered by magnitude to reduce a set of *p* correlated variables to a smaller set of *k < p* orthogonal variables. Versions of PCA for spatio-temporal data were referred to by,^19^ S-mode and T-mode PCA, the particular mode depending on whether the columns of are time points (T-mode) or countries (S-mode). The S-Mode PCA aims to find dominant temporal trends across the spatial locations, highlighting a small number of dominant temporal trends across all countries. Conversely, T-Mode PCA aims to find different spatial patterns in the data and the associated time points at which they occur. In general, however, PCA finds unsupervised structures in the data by conducting an eigen decomposition of the correlation of covariance matrix. Whilst this can often be useful in visualising data in lower dimensions, it is not possible to guide the structure of the new axes using prior information or independent data. In order to account for known spatio-temporal correlations inherent in the data, we use spatio-temporally weighted S-mode and T-mode PCA, which aim to find dominant temporal and spatial patterns respectively.^7^ extended these approaches to account for known spatio-temporal structures in river flow systems through the use of spatial and/or temporal weight matrices to inform spatio-temporal structure. Assuming the data matrix is a *n × p* matrix, a *p × p* column weight matrix **Ω** and *n × n* row weight matrix **Φ** can be constructed so that PCA is applied to a transformed matrix *X′* = **Φ*X*Ω**. We adopt a similar approach to considering the transmission of COVID-19 through air travel as a proxy to general movement between countries. Residual spatial or temporal structure may then be associated with the impact of any intervention method introduced in that country. Analyses were conducted using the stpca package in R.

#### 3.4.1 Spatial weight matrix

Using data on air traffic between airports in the different countries, we construct a spatial weight matrix using a similar approach to.^7^ The method involves constructing a spatial weight matrix of air passenger connectivity between countries for which COVID-19 case and death data were available. The air passenger data provided by^20^ generated a modeled passenger flow matrix for all airports with a host city-population of more than 100,000 and within two transfers of air travel from various publicly available air travel datasets. Multiple covariates were included in a spatial interaction framework to predict the air transportation flows between airports. This modelled flow matrix is used here to explain flow of passengers between different international airports, much in the same way that hydrological flow is measured in.^21^

Initially, all starting airports and destination airports were assigned to a country using the GNcountryCode function in the geonames package in R, based on their latitude and longitude. These were then matched to the corresponding countries in the case and death data. The entries of spatial weight matrix *S*_*i,j*_ denoting the flow of passengers into country *j* from country *i* were then the sum of all individual flows of passengers to/from airports within that country. To ensure valid covariance matrices, diagonal terms are kept equal to 1 and the remaining terms are scaled by the row means of the remaining non-diagonal terms.

Once this initial matrix has been calculated, to ensure a valid postive-definite covariance matrix is produced, the final weight matrix is calculated as

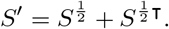

where 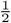 is the matrix squareroot and denotes the matrix transpose.

#### 3.4.2 Temporal weight matrix

The temporal weight matrix is constructed similarly to.^7^ Independent Generalised Additive Modesls (GAMs)^22^ are fitted by restricted maximum likelihood (REML) in the mgcv package^23^ to each of the time series with an intercept and an univariate smooth function of date since 1st January 2020 as predictor. Remaining correlation in the model residuals between time [1, …, (*n* − 1)] and [2, …, *n*] is calculated for each country and then the median value is used as an estimate of *ρ*, the average global temporal correlation. This aims to remove general seasonal patterns for each country that can be accounted for by simple smooth functions of time, before attributing latent residual correlation to additional epidemiological. Joint latent covariance models are frequently used to detect or account for residual joint covariance across a variety of applications.^24^ The median residual temporal correlations were estimated as 0.812 and 0.808 for cases and deaths respectively. The *i*th row and *j*th column element of the temporal weight matrix *T*_*i,j*_ is then specified as *ρ*^|*i−j*|^ for all time indices in the original data matrix.

## 4 Conclusions

Our results show a strong temporal pattern in both the global case and death numbers, with comparatively minimal spatial pattern. Only a single principal component is required in each case to explain the vast majority of variation in the data, suggesting consistent dominant temporal trends ocross the globe. Previous analyses of global data have focused on fitting models independently for each country and compared results post analysis^26^, whereas our approaches model all countries simultaneously through time. The two approaches were complimentary, rather than contradictory. Aruba was consistently found to show different dynamics to other countries and was selected as a unique cluster in the clustering approaches and was also singled out in the T-mode PCA analyses. The data for Aruba is plotted in Figure 9.

**Figure 9:**
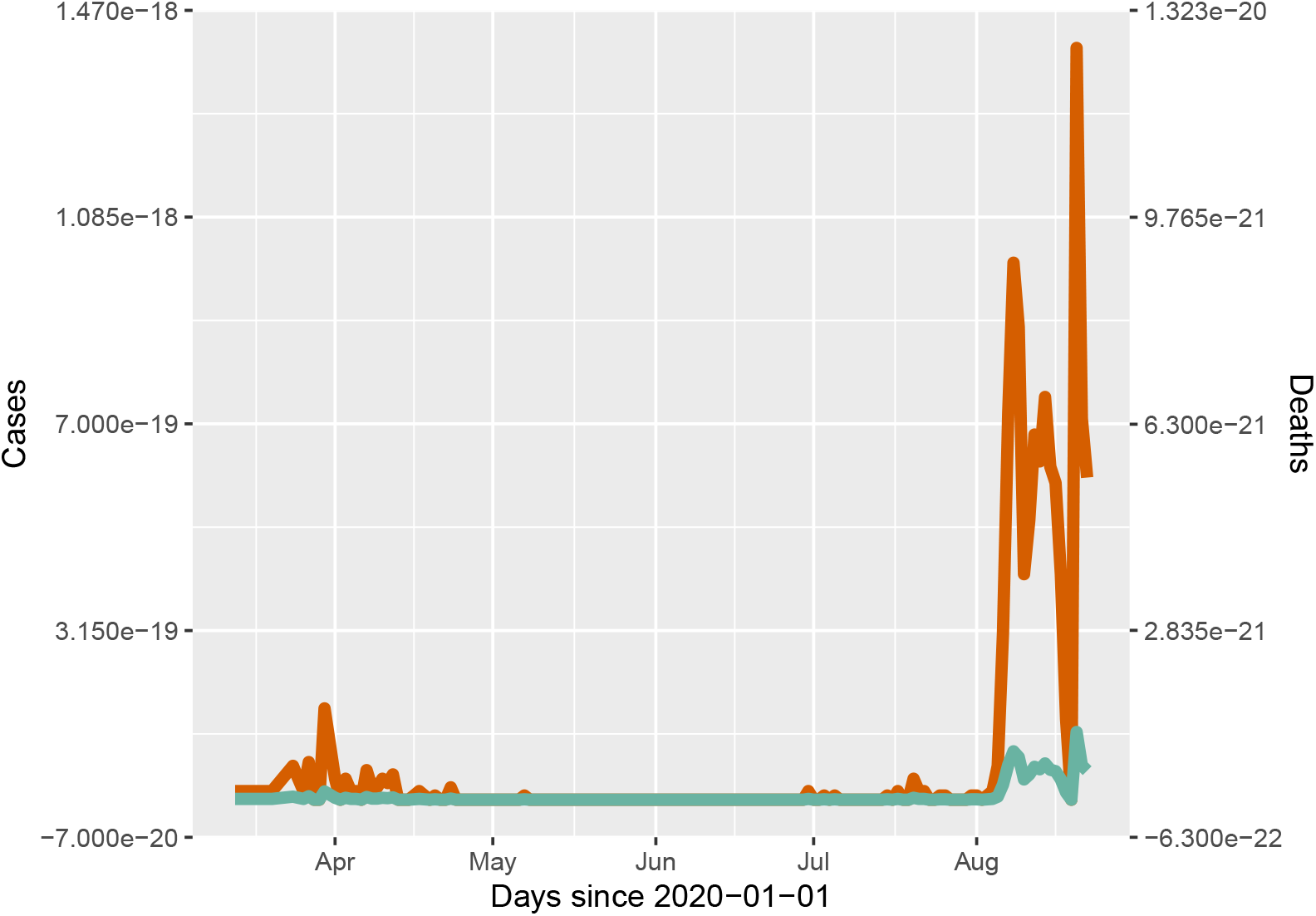
Cases (red, left vertical axis) and deaths (green, right vertical axis) per capita in Aruba.

The clustering according to temporal correlation in the data has the benefit of being able to use full trajectories from the beginning of January and allows all countries with data at any point during the pandemic to be included. This approch is particularly successful at joining together similar temporal trajectories, avoiding the separation of countries that have similar dynamics but larger populations and hence larger absolute number of cases. The clustering also suggests that temporal correlation in both cases and deaths is significant even at lags of up to 20 days for some countries, whilst others have spikes resulting in low correlation for all lags greater than two.

Partition clustering according to numbers of cases and deaths is successful at clustering temporal trajectories, but once again there was little similarity between clusterings according to case numbers and those conducted on mortality data.

The weighted PCA shows that there is important spatial and temporal structure within the data, and when this is removed, many of the important joint global trends are more easily detected. The temporal dynamics are shown to be particularly important in the global spread of infected individuals. Providing a semi-supervised approach, the method more readily extracts joint structure and highlights the emergence of a spike in numbers of cases globally from early August, with a subsequent spike in deaths approximately a week later. This appears consistent with the increase in mutant strains observed in global studies.^28^ There are two obvious daily spikes in the S-Mode analysis of deaths that is driving the principal temporal trends, and this may well correspond to a lag in data collection or data dumping. Subsequent principal components showed the complementary pattern to those observed in the case data.

As such, this approach offers the possibility of real-time detection of changes in pandemic dynamics through extraction of important changepoints, where analyses are re-run when new data are collected, which may correspond to the emergence of new strains with contrasting properties or issues with data collection protocols that require further study. The approach is incredibly fast computationally and easily highlights important and interpretable trends in complex and potentially very noisy spatio-temporal data. These can then formally assist with policy decisions in relation to when and where interventions appear to be working^26^ and when they are not, which^2^ linked to cultural tightness. The emergence of significant changes in dynamics can also trigger further epidemiological and genetic studies to detect viral mutations that may be associated with or causing the changing dynamics.

## 5 Data availability

All data are publicly available and references and access dates are provided within the text.

## Data Availability

All data are publicly available and references and access dates are provided within the text.

https://www.ecdc.europa.eu/en/publications-data/download-todays-data-geographic-distribution-covid-19-cases-worldwide

https://data.worldbank.org/indicator/SP.POP.TOTL

https://www.worldpop.org/project/categories?id=13

